# ULTRASOUND OF QUADRICEPS FEMORIS AS A RULE-OUT TOOL FOR MALNUTRITION SCREENING IN HEART FAILURE CLINICS

**DOI:** 10.64898/2026.09.02.26362047

**Authors:** Paula A B Ribeiro, Priccila Zuchinalli, Zoe Carrier, Olivier Nguyen, Anne-Sophie Zenses, Gabriela Corra Souza, François Tournoux

## Abstract

**Aims:** To assess reproducibility and repeatability of quadriceps femoris muscle (QFM) thickness measured by 2D ultrasound (US) and evaluate its potential as a tool to detect risk of malnutrition in heart failure (HF) outpatients.

**Methods:** Thirty HF participants (malnourished n=16; well-nourished n=14) were recruited. Malnutrition was assessed using the GLIM tool (reference method). QFM thickness was measured at two points (mid-point and 2/3-point). We conducted paired T-tests to assess differences in QFM thickness according to leg side (left vs. right), condition of measure (with vs. without probe pressure), inter- and intra-operators. Intraclass coefficients (ICC) were used to test intra- and inter-operator agreement.

**Results:** Analyses showed no differences for most comparisons (total of 48). The midpoint was different in 4 comparisons, 2/3 point for none. ICC analyses ranged from 0.943 to 0.996 for intra- and inter-operator analyses. The best result from the ROC analyses was for the 2/3 QFM point with no probe pressure (AUC 0.74), with a specificity of 43% and sensitivity of 100%.

**Conclusion:** Our preliminary results suggest that 2D – US QFM thickness, measured at the two-thirds point without pressure, is a reproducible and repeatable measurement. QFM thickness also showed good ability to differentiate between nutritional statuses and to rule out malnutrition (high sensitivity). These findings indicate it as a practical initial screening tool for identifying patients who do not need further investigation. However, these preliminary findings need to be confirmed in a larger sample of patients.

## INTRODUCTION

Patients’ nutritional status is a key component of overall health and a well-established predictor of clinical outcomes in cardiac patients^1-4^. In heart failure (HF), poor nutritional status and wasting have been reported across the spectrum of both preserved and reduced ejection fraction, with a multifactorial etiology^5-8^. Malnutrition is associated with increased morbidity and mortality, impaired quality of life, and higher healthcare costs^9^. Despite its high reported prevalence in HF—ranging from 16% to 90% depending on the population and assessment method—malnutrition is not systematically assessed in specialized HF clinics.

Several tools and criteria exist to identify malnutrition in the general population; however, no single approach has achieved universal acceptance. To address the lack of a gold standard, the Global Leadership Initiative on Malnutrition (GLIM) proposed diagnostic criteria combining phenotypic and etiologic components^10^. While GLIM represents an important advance, its implementation in routine outpatient HF clinics is limited by time constraints and training requirements, which likely contributes to the persistent underdiagnosis of malnutrition in this population.

Muscle mass (MM) status is directly related to malnutrition diagnosis and severity staging. Loss of MM is independently associated with reduced quality of life, increased susceptibility to infections, frailty, and higher mortality risk^4^. In clinical practice, malnutrition is often identified only at advanced stages, when frailty or cachexia is already established. Although multiple techniques are available to assess MM, their use is frequently dictated by local availability, feasibility, and the need for population-specific cut-off values. Emerging evidence suggests that ultrasonography (US) is a reliable, rapid, and cost-effective method for MM assessment across various clinical settings, particularly in critically ill populations^7,11^, and has been validated in oedematous patients—an important advantage in HF.

Given the limited feasibility of nutritional assessments in high-volume and high malnutrition prevalence in outpatient HF clinics, there is a clear unmet need for a simple and accurate screening tool to identify patients at risk of malnutrition and prompt early intervention. We therefore hypothesized that practical barriers faced by healthcare professionals contribute to malnutrition remaining an underrecognized morbidity in HF care.

In this context, our study aimed to evaluate the reproducibility and repeatability of ultrasonographic measurement of quadriceps femoris muscle (QFM) thickness, and to compare patient groups according to GLIM criteria. We further sought to determine its discriminatory potential as a pre-screening tool for identifying HF outpatients who require a comprehensive nutritional assessment.

## Methods

### Patients recruitment

Patients from the HF specialized outpatient clinic and hospitalization with a diagnosis of chronic HF were recruited according to their nutritional status. We estimate that at least 50% of our patients are malnourished, therefore, they were recruited in a proportion of 1/1 of malnourished to well-nourished until we achieved 30 patients.

### Exclusion criteria

Physical conditions that prevent the operator from performing the QFM measure, such as amputation, patients using a wheelchair, disabilities that make it impossible for the patient to maintain the position necessary for evaluation; Patients with known inflammatory muscle disease; and patients who were not able to consent.

### Data collection

Clinical data was collected from electronic medical records (i.e., NYHA class, comorbidities, echocardiogram parameters, blood markers and medication).

### GLIM assessment

The Global Leadership Initiative on Malnutrition (GLIM)^10^ instrument was applied by a trained clinical research nurse to detect malnutrition. The instrument requires at least one phenotypic criterion and one etiologic criterion for diagnosis. Figure 1 presents how we applied the criteria in our study:

**Figure 1.**
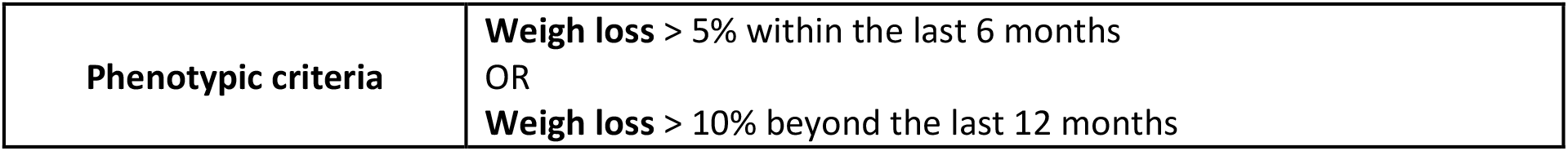
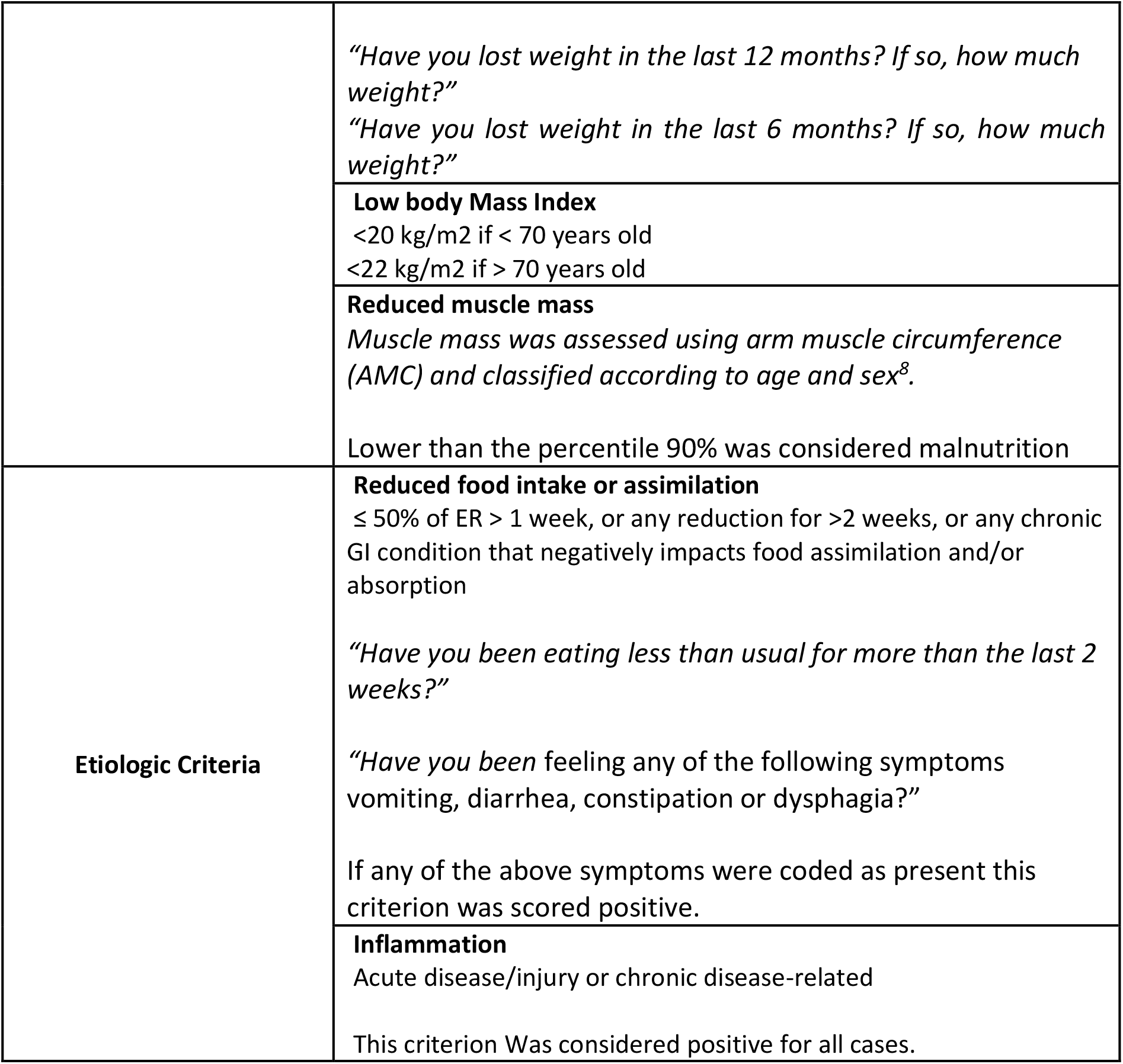
Criteria and applied method for GLIM assessment during clinical routine.

**Figure 1.**
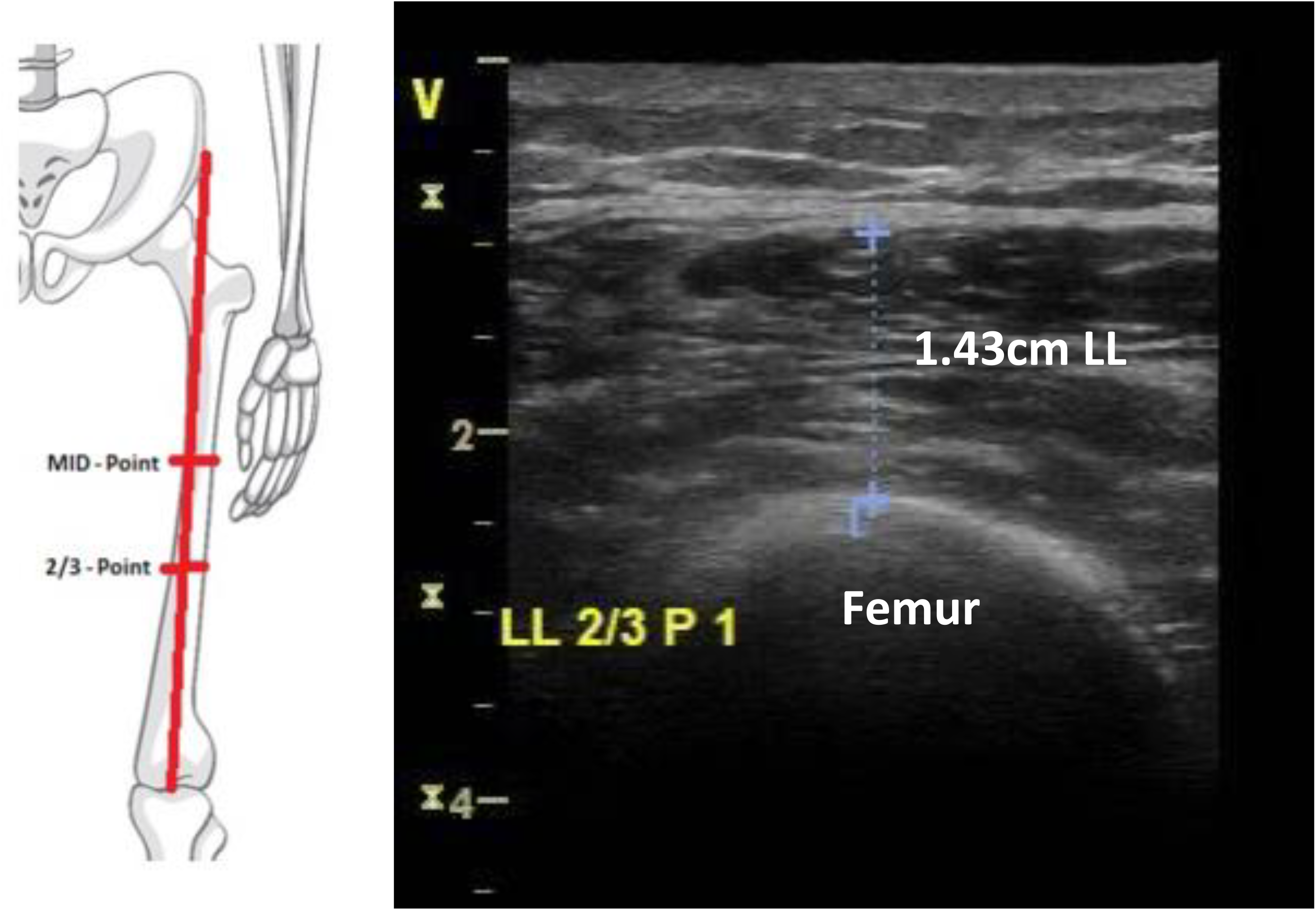
Anatomical diagram and a transverse ultrasound representative measure of the quadricep femoris muscle (left leg) – pressure condition.

**Figure 2.**
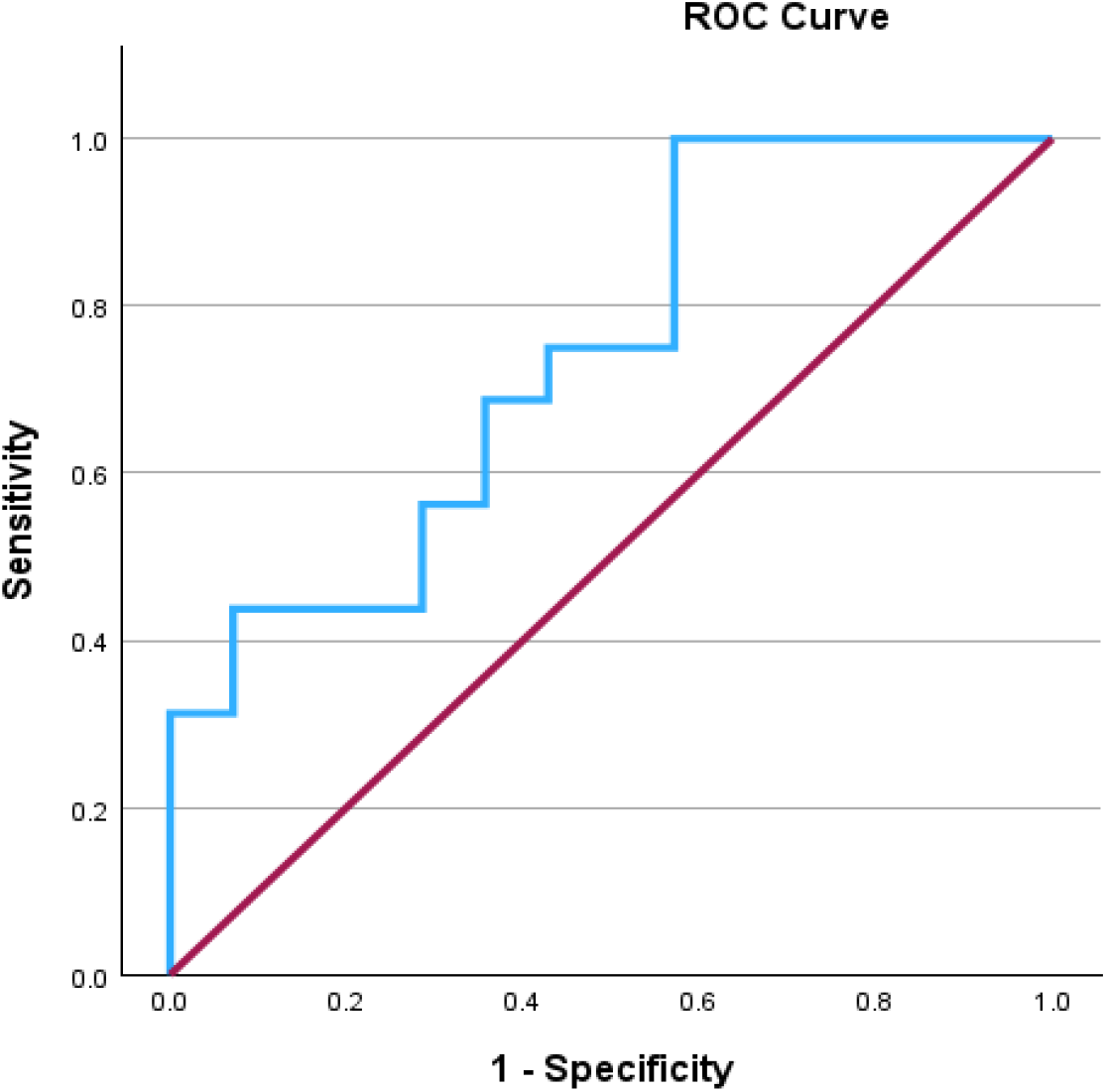
QFM thickness according to pressure condition and GLIM status.

### QFM ultrasound images: data acquisition

Two trained health care professionals performed the measure of the QFM thickness. QFM images were acquired using GE Vivid S6 2D US (GE Medical System, Milwaukee, WI, USA), equipped with a 13 MHz transducer. Acquisitions were performed with the patient in lying position and extended legs, in a muscle relaxed state. All QFM measurements were performed by at least two healthcare care professionals from our team (i.e. clinical nurse, a nutritionist and a medical student). Thickness refers to the width of the QF muscle (without fat tissue) and was calculated using a perpendicular line to the horizontal axis from the point of the femur to the muscle fascia. The QFM was determined at the mid-point and 2/3-point between the anterior superior iliac spine and the upper border of the patella, from the right and the left leg, with and without probe compression. Imaging was measured at the moment of acquisition, and all measures were performed twice per evaluator.

### Data processing and statistical analysis

Continuous data are presented in mean ± standard deviation (SD). Categorical data are presented in absolute (n) and relative frequency (%). Intra-class coefficient (ICC) was performed to analyse inter-observer and intra-rate agreement. Paired Student’s T-test was used to analyse the mean differences of the QFM thickness value between well nourished and malnourished according to GLIM groups. All statistical tests were conducted with the SPSS statistics software, version 28. The significance level will be set at p≤0.05 for all analyses.

## Results

Patients were recruited from March to December 2022. Clinical characteristics are described in Table 1 according to GLIM status (i.e., well-nourished vs. malnourished). For that, we merged moderate malnutrition (n=12) with severe malnutrition (n=3) and created two groups. Patients were different for the NYHA distribution according to GLIM status, and mid-arm muscle circumference (details in Table 1).

**Table 1.** Clinical characteristics according to the GLIM nutritional status.

| GLIM STATUS |  |  |  |  |
| --- | --- | --- | --- | --- |
|  | Well nourished<br>(n=14) | Malnourished<br>(n=16) | TOTAL<br>(n=30) | p |
| Age | 65.6 ± 13.2 | 69.6 ± 13.4 | 67.7 ± 13.2 | 0.425 |
| Gender (M/F) | 10/4 | 11/5 | 21/9 | 0.873 |
| BMI | 30.0 ± 5.6 | 28.4 ± 8.1 | 29.1 ± 7.5 | 0.553 |
| **NYHA** |  |  |  |  |
| I | 6 (40.0) | 0 (0) | 6 (20.0) | **0.009** |
| II | 5 (33.3) | 7 (46.7) | 12 (40.0) |  |
| III | 0 (0) | 6 (40.0) | 6 (20.0) |  |
| IV | 1 (6.7) | 0 (0) | 1 (3.3) |  |
| Unknown/unclassified | 3 (20.0) | 2 (13.3) | 5 (16.7) |  |
| LVEF (%) | 47.3 ± 15.0 | 40.2 ± 15.4 | 43.4 ± 15.4 | 0.220 |
| LVEF reduced (<45%) | 5 (38.5) | 8 (50.0) | 13 (44.8) | 0.534 |
| **Self-report GI symptoms** |  |  |  |  |
| Nausea or vomiting | 1 (7.1) | 1 (6.3) | 2 (6.7) | 0.922 |
| Diarrhea or constipation | 4 (28.6) | 4 (25.0) | 8 (26.7) | 0.825 |
| Gastroparesis | 2 (14.3) | 3 (18.8) | 5 (16.7) | 0.743 |
| Self-report weight loss last year | 1 (7.1) | 3 (18.8) | 4 (13.3) | 0.351 |
| Weight loss (range %) | 2.5 | 9.1 – 14.0 | 2.5 – 14.0 | 0.098 |
| Diabetes type II n (%) | 5 (35.7) | 6 (37.5) | 11 (36.7) | 0.919 |
| Hypertension n (%) | 10 (71.4) | 9 (56.3) | 19 (63.3) | 0.389 |
| Chronic kidney disease | 6 (42.9) | 7 (43.8) | 13 (43.3) | 0.961 |
| **Medication use – n (%)** |  |  |  |  |
| Beta-blocker (n=30) | 11 (78.6) | 15 (93.8) | 26 | 0.222 |
| ACE OR ARB (n=30) | 6 (42.9) | 4 (25.0) | 10 | 0.301 |
| Amiodarone (n=29) | 1 (7.7) | 2 (12.5) | 3 | 0.672 |
| Warfarin (n=29) | 3 (23.1) | 0 (0) | 3 | **0.042** |
| NOAC (n=29) | 3 (21.4) | 6 (40.0) | 9 | 0.280 |
| Digoxin (n=30) | 1 (7.1) | 2 (12.5) | 3 | 0.626 |
| Diuretic (n=30) | 8 (57.1) | 10 (62.5) | 18 | 0.765 |
| Spironolactone (n=30) | 4 (28.6) | 12 (75.0) | 16 | **0.011** |
| CACB (n=28) | 1 (7.7) | 2 (13.3) | 3 | 0.630 |
| Entresto (n=30) | 4 (28.6) | 8 (50.0) | 12 | 0.232 |
| SGLT2 (n=30) | 3 (21.4) | 9 (56.3) | 12 | 0.052 |
| Nt pro BNP (n13 vs 15) | 1901 ± 2714 | 4491 ± 8772 | 3289 ± 6701 | 0.317 |
| Lymphocytes (n11 vs 12) | 2.5 ± 2.4 | 1.5 ± 0.73 | 1.97 ± 1.78 | 0.221 |
| **Anthropometric parameters** |  |  |  |  |
| Mid-arm muscle circumference | 25.9 ± 3.4 | 20.3 ± 2.9 | 22.9 ± 4.2 | **<0.001** |
|  | Well nourished<br>(n=14) | Malnourished<br>(n=16) | TOTAL<br>(n=30) | p |
| Arm circumference | 31.3 ± 5.3 | 27.9 ± 4.9 | 29.5 ± 5.3 | 0.076 |
| Triceps skinfold | 17.9 ± 12.6 | 23.3 ± 11.3 | 20.8 ± 12.1 | 0.222 |

GLIM: Global Leadership Initiative on Malnutrition; Values are presented as mean ± SD or numbers of patients (percentages); p for Student T test for independent samples.

Table 2 presents the means, SD, and ICC for intra- and inter-operator measurement means, both legs (left and right), and with and without probe pressure conditions. T-test analyses demonstrated no differences for most comparisons, except mid-point QFM right and left legs – no pressure condition and mid-point QFM right leg – pressure condition (both intra evaluator); and also, mid-point, right leg - no pressure condition (inter operator). The right vs left leg showed a difference for the mid-point for one of the operators, both rounds. The 2/3 thigh point showed no difference for any of the comparisons. Regardless, overall agreement analyses (ICC) were high in all comparisons. Overall, ICC ranged from 0.942 to 0.996 for the intra-operator comparisons, and from 0.894 to 0.977 for inter-operator comparisons.

**Table 2.**
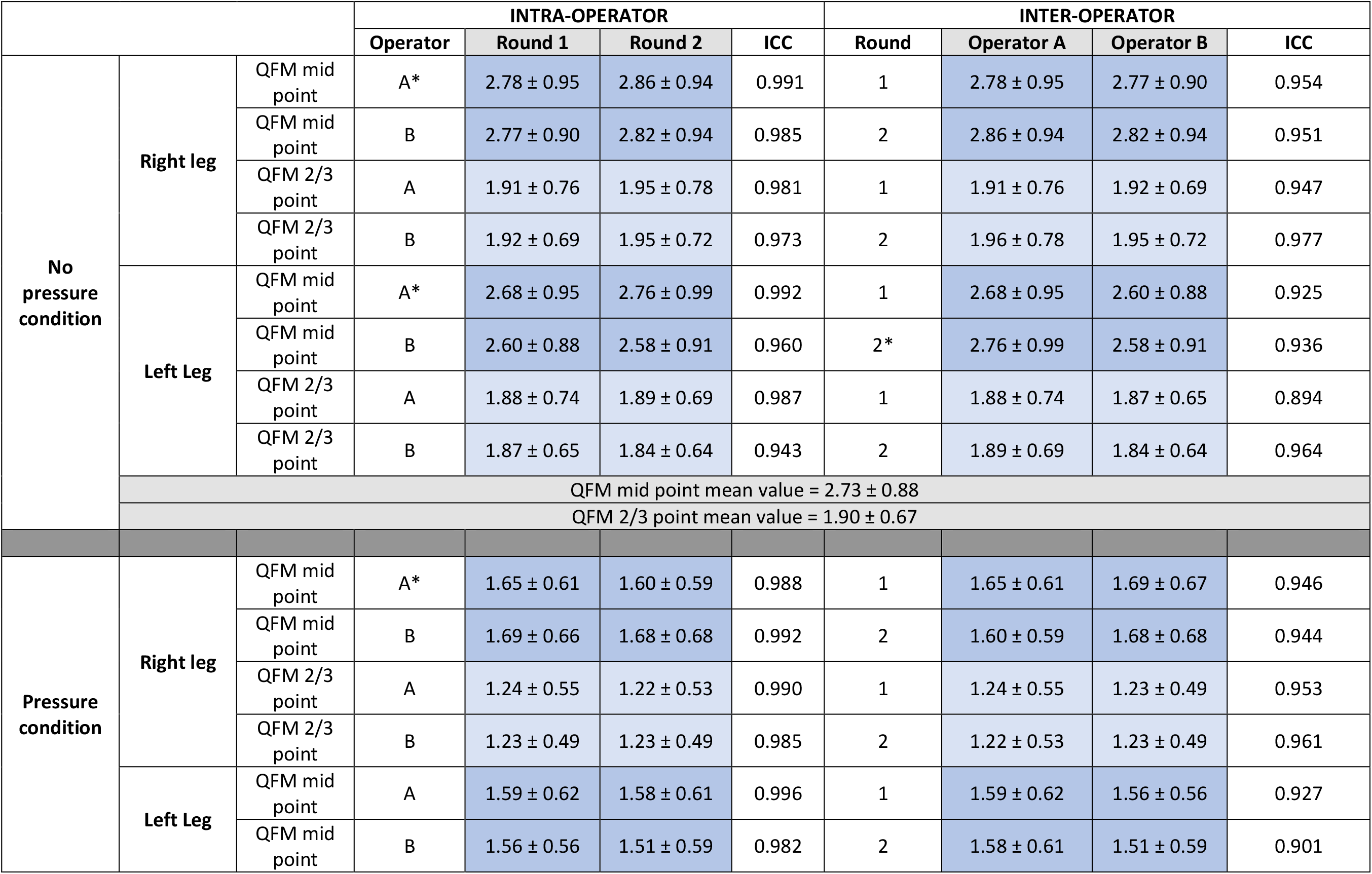
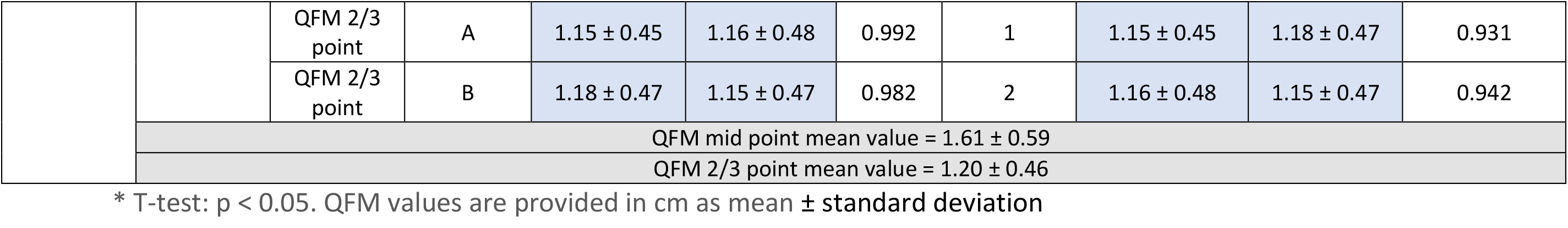
Comparisons of QFM thicknesses within and between operators in all tested conditions.

Figure 3 presents the results for AUC for 2/3 QFM point for pressure (panel A) and no pressure (panel B). ROC analysis was significant for the 2/3 QFM point (p=0.027 and p=0.008) for both pressure and no pressure conditions, respectively.

AUC for the 2/3 QFM point pressure condition was 0.710 and demonstrated that the cut-off point of 1.01 cm has 50% sensitivity and 85.7% specificity. When no pressure condition was analyzed, AUC for 2/3 QFM point was 0.741, with a cut-off point of 2.43 and 100% sensitivity, and 43% specificity. In practice, in our sample, eight patients classified as well nourished according to GLIM had QFM < 2.43; however, 0 out of 16 patients in the malnourished group had QFM > 2.43.

## Discussion

Our study shows preliminary results demonstrating QFM thickness measured by US seems to be a good tool for pre-screening malnutrition in HF outpatients. QFM thickness at the 2/3-point showed better reproducibility and repeatability of measurements compared to mid-point measurements. Further, a larger area under the curve was observed when tested for malnutrition diagnosis, when compared to the QFM mid-point. Concerning the pressure condition, no probe pressure performed better than with pressure, contrary to what was previously described^12^. QFM also demonstrated that it is inversely correlated with an important HF biomarker – NT-proBNP. Altogether, we believe it is a promising tool with easy implementation in the clinical routine. The 2.43cm cut-off was able to exclude malnutrition among these patients, therefore an interesting rule-out tool. Also, interestingly, none of the malnourished patients were NYHA class 1, meaning that malnourishment might have a relationship with disease severity in these patients.

The results demonstrated high ICC intra and inter-evaluators. Our results are in agreement with previous studies^110^ that demonstrated US reliability in healthy^7^, CAD patients^13^, and bedside ICU patients.^14,15^ The acquiition technique is simple, allowing healthcare professionals to easily evaluate in the clinical routine, regardless of specific US training - none of our evaluators had previous formal imaging acquisition training. In general, high-resolution ultrasounds make it possible to visualize the dermis and subcutaneous layers (i.e., adipocyte and connective tissues), and muscle tissue as a hypoechogenic layer, making measurement an easy and quick task.

A recent systematic review^11^ described the current use of muscle US tools to predict clinical outcomes (i.e., intensive care unit-acquired weakness, nutritional status, and length of hospital stay). The quadriceps femoris muscles were the most studied muscle group due to their accessibility and size. Most literature focuses on ICU patients (22 out of 37 studies), and there is still significant heterogeneity in terms of ultrasound protocol and outcomes measurements across studies. Importantly, only one study tried to correlate with biomarkers, and five studies with nutritional status. Also, previous studies demonstrated that the rectus femoris diameter is a good clinical marker and is associated with functional capacity and VO_2_max^6,8,11^ in HF patients.

Moreover, surprisingly, a follow-up study suggested the US muscle thickness assessment is a reliable method to detect changes over time (i.e. short-term changes), while GLIM was not, suggesting that US measurements could be a better tool^16^. In the clinical routine, changes in QFM are not expected in the short term, unless patients are hospitalized. Considering the re-hospitalization rates are 20 to 25% in 30 days,^17^ it is not unusual that HF patients spend periods of time with reduced mobility and bed rest; it could be the easiest tool to implement when a surveillance system is necessary (i.e., post-hospitalization, long term follow-ups, major weight change, etc.).

## Limitations

Our small sample size that doesn’t allow us to explore the comparisons of specific subgroups, such as LVEF reduced vs. preserved, BMI status, or sex differences. We are aware that our results need to be replicated in a larger sample and in a long-term fashion to verify how well it performs in detecting changes over time. Still, we believe our study is a first step, from a cross-sectional perspective, before testing sensitivity to changes in muscle status.

## CONCLUSION

Our preliminary results suggest that QFM thickness, measured using two-dimensional ultrasound without applied pressure, is a reproducible and repeatable parameter. Measurements performed at the two-thirds point demonstrated better performance than those obtained at the mid-point. QFM thickness also showed good ability to differentiate between nutritional statuses and to rule out malnutrition in our study population (high sensitivity). These findings indicate that ultrasonographic assessment of QFM thickness may represent a practical initial screening tool for identifying patients who do not need further investigation. The next phase of this work will aim to confirm these preliminary results in a larger cohort and to evaluate the performance of this method in detecting longitudinal changes in nutritional status over time.

## Data Availability

All data produced in the present study are available upon reasonable request to the authors.

## Acknowledgements

We sincerely thank all the patients who participated in this study. We are deeply grateful for their time, trust, and willingness to contribute to clinical research.

